# Time course of Quadriceps thickness changes over six months post-anterior cruciate ligament reconstruction: unveiling critical impairments in vastii muscles

**DOI:** 10.1101/2024.11.05.24316670

**Authors:** Carlos De la Fuente, Rony Silvestre, Eduardo Martinez-Valdes, Roberto Yañez, Ferran Abat, Alejandro Neira, André Gustavo de Andrade, Felipe P Carpes

## Abstract

The mechanisms behind the persistent Quadriceps weakness observed after an Anterior Cruciate Ligament reconstruction (ACLr) remain debatable. We described the time course of changes in Quadriceps thickness, strength, and thigh circumference over six months following an ACLr. We also studied the causal relation between Quadriceps thickness, total thickness and strength. Quadriceps thicknesses, Quadriceps strength, and thigh circumference were measured preoperative, 3, and 6 months post-ACLr surgery in 103 patients (77 men and 26 women). Limbs and time were compared with repeated-measures ANOVA and causal relations through mediation analysis (α = 5%). From 0 to 3 months post-surgery, Quadriceps strength (p<0.05), and VI (p<0.05) and VL (p<0.001) thicknesses was reduced. From 3 to 6 months, Quadriceps strength (p<0.05), total Quadriceps thickness (p<0.001), and VI (p<0.001) and VL (p<0.05) thicknesses increased. VM and RF thicknesses increase to 3 months after ACLr (p<0.05). Quadriceps strength, circumference at 5 cm, VI, VL, VM, and total Quadriceps thickness remain lower than the contralateral limb 6 months following an ACLr (p<0.05). The VI thickness determine indirectly (p<0.001) through the total Quadriceps thickness the Quadriceps strength pattern, while the VL thickness directly determined it (p=0.034). Quadriceps thickness adapts heterogeneously, with VI and VL atrophying for up to three months, explaining the post-ACLr quadriceps weakness. By 6 months, VI does not recover its thickness proportion, and VI and VL do not return to pre-surgery conditions. Both vastii muscles primarily determine Quadriceps strength changes, eliciting the VI thickness as a key biomarker for Quadriceps weakness following ACLr.

## 1. Introduction

Quadriceps muscle weakness can last up to 12 months following an Anterior Cruciate Ligament reconstruction (ACLr) (Czaplicki et al., 2015; Pottkotter et al., 2020). Quadriceps weakness leads to poor patient outcomes, early onset of osteoarthritis, and increased ACL re-rupture risk (Criss et al., 2023). The etiopathogenesis of muscle weakness is primarily a condition of muscle atrophy (Birchmeier et al., 2020; Norte et al., 2018; Tim-Yun Ong et al., 2022), but neural and inhibitory circulant molecules can also promote persistent muscle weakness (De la Fuente et al., 2022b; Fukunaga et al., 2019; Hennebry et al., 2017; Kuenze et al., 2015; Nuccio et al., 2021; Pietrosimone et al., 2022; Yang et al., 2019). Although Quadriceps muscles may re-store their morphological characteristics post-ACLr, heterogeneous changes between the different Quadriceps muscle heads may induce alterations in muscle synergies and overall deficits in Quadriceps strength. Unfortunately, there is scarce evidence regarding the adaptation of the different Quadriceps muscle heads and its impact on Quadriceps strength after ACLr. Thus, changes in Quadriceps thickness, a technique with moderate to high reliability (ICCs from 0.76 to 0.95 (Abiko et al., 2022)), may serve as a clinically useful tool to potentially assess muscle weakness (Harput et al., 2022; Lanferdini et al., 2023). Similarly, this method has been previously explored for assessing other muscle dysfunctions (De la Fuente et al., 2020).

A reduction in vastus intermedius (VI) and vastus lateralis (VL) muscle thickness is observed within 48 to 72 hours in the early postoperative phase (Lee et al., 2020). Seven days later, all Quadriceps portions show a reduction in muscle thickness, accompanied by lower levels of IGF-1 and a slight increase in myostatin levels (Yang et al., 2019). By week six following ACLr, functional outcomes from the International Knee Documentation Committee (IKDC) (Irrgang et al., 2001) begin to improve (Agarwal et al., 2021). Three months following ACLr, changes in Quadriceps volume (total volume) can explain 57% of Quadriceps strength variance (Grapar Žargi et al., 2017). Following the first six postoperative months, Quadriceps strength deficits appear more closely associated with Quadriceps morphology (Pottkotter et al., 2020; Thomas et al., 2016) rather than neural inhibition (Fukunaga et al., 2019; Harput et al., 2022). Thus, the understanding of differences in muscle thickness adaptations following ACLr is relevant not only for clinical purposes of understanding the possible sources of weakness and better planning exercise therapies based on dysfunctions to different Quadriceps muscle heads (Cobian et al., 2024) but also is relevant for further studies addressing fatigue effects (Bouillard et al., 2014) or force output interpretations like isometric and isokinetic measures following ACLr (Bouillard et al., 2014).

Understanding Quadriceps thickness adaptations following ACLr is important to distinguish morphological causes of Quadriceps weakness (De la Fuente et al., 2022b; Pietrosimone et al., 2022; Sonnery-Cottet et al., 2019). Although the Quadriceps thickness and circumference measurements may show deficits between 5% and 23% compared to the contralateral limb (Agarwal et al., 2021), patients demonstrate strength and functional clinical improvements six months post surgery (Pottkotter et al., 2020; Thomas et al., 2016). In consequence, understanding how Quadriceps thickness changes across all of its muscle heads and determining how knee extension strength, and thigh circumference change during the initial six months after ACLr, could provide important information regarding rehabilitation progression. Furthermore, establishing the causal relation between Quadriceps thickness, total thickness, and Quadriceps strength during this period, could provide specific therapeutic targets for the management of the injury. This understanding may unveil unknown Quadriceps morphology changes within the first six months of evolution and markers of Quadriceps weakness.

Therefore, in this study, we set out to determine whether all Quadriceps muscles adapt similarly during the six months following an ACLr that leads to muscle weakness. Furthermore, we determine the causal relation between Quadriceps thickness, total thickness, and Quadriceps strength in ACLr patients over the six months of evolution post surgery.

### 2. Patients and Methods

#### 2.1. Study design

In this retrospective, observational, and comparative study, patients with a first acute and unilateral ACLr were evaluated for Quadriceps thickness, thigh circumference, and Quadriceps strength from pre-surgery up to six months after an ACLRr. Measurements were performed in the ACLr and contralateral leg [CON] at 3-time points (pre-surgery [0M] without anesthesia, three months post-surgery [3M], and six months post-surgery [6M]). Time points and groups were compared. Two experienced physical therapists trained in ultrasonography (> 5 years of experience) collected all the data.

#### 2.2. Participants

From February 2021 to April 2024, 103 adult patients with arthroscopic ACLr (77 men and 26 women; age 29.0 ± 10.2 years, height 1.73 ± 0.07 m, body mass 77.2 ± 11.1 kg, body mass index 26.0 ± 3.8 kg/m^2^, pre-operative IKDC 51.5 ± 16.4 pts, IKDC at three months 58.6 ± 13.8 pts, and IKDC at six months 74.3 ± 12.5 pts) were included in this study. The inclusion criteria were: i) first unilateral acute ACL rupture, ii) positive Lachman and anterior drawer test, iii) non-contact sports-related ACL rupture, iv) arthroscopic ACL complete mid-substance rupture confirmation, and v) age between 18 and 55 years old. The exclusion criteria were any rheumatologic, neurologic, and metabolic conditions. The local institution’s ethics committee of MEDS clinic (#122024) approved this research, which was conducted according to the Helsinki Declaration, and written informed consent was given.

#### 2.3. Diagnosis and rehabilitation

A combination of physical examinations associated with acute inflammation signs, positive anterior drawer tests, and non-integrity of the ACL assessed by magnetic resonance imaging (MRI) served for the ACL rupture diagnosis hypothesis, which was confirmed by arthroscopy (Caracciolo et al., 2021; Yañez-Diaz et al., 2023). The MRI showed focal or diffuse discontinuity of the ACL with a preference for sagittal fast spin-echo T2, abnormal ACL signal (focal or diffuse increased signal intensity within discontinuous ACL) with a preference for the sagittal fast spin-echo T2-fat-suppressed, the decreased slope of residual ACL fibers, and a wavy configuration of the ACL with an angle lower than 45° (ACL–Blumensaat line) with a preference in the sagittal oblique fast spin-echo T2 (Caracciolo et al., 2021; Yañez-Diaz et al., 2023).

All patients completed a six-month physiotherapy intervention following the same protocol (San Martín-Mohr et al., 2018). The goals for the first six weeks of rehabilitation were to reduce inflammation, restore the knee range of motion, favor muscle activation with open kinetic chain exercises (Boccia et al., 2019), gait training using crutches (De la Fuente et al., 2022a), and control excessive loads considering both the type of graft and patient conditions. Knee strengthening and neuromuscular control exercises started six weeks post-surgery (San Martín-Mohr et al., 2018). Symmetrical knee strength and jogging started 12 weeks post-surgery (San Martín-Mohr et al., 2018). Plyometric exercises, jumping, and change of direction movements in close kinetic chain exercises started 20 weeks post-surgery (San Martín-Mohr et al., 2018).

#### 2.4. Quadriceps thickness assessment

The thickness of the Quadriceps was measured by physical therapy with 5 yrs of experience measuring at each time point ([0M], [3M], and [6M]) by ultrasonography images acquired using a handheld Lumify ultrasound system (Philips Ultrasound, Inc., The Netherlands) in B-mode, 34.5 mm field of view, linear array transducer L12-4 (Philips Ultrasound, Inc., The Netherlands), and probe frequency of 12 MHz. Images were sampled at 28 Hz, TLB of 0.1, Musculoskeletal pre-configuration, power of −0.3 dB, adjusted gain between 40 and 60 points, and depth between 4.0 and 6.0 cm depending on conditions of participants, i.e., greater subcutaneous tissue thickness. Before the acquisition, the patients rested for at least 10 min (Lanferdini et al., 2023) in a comfortable room at 22 °C. A water-soluble gel was applied in the ultrasonographic probe to provide complete acoustic contact with the skin (Lanferdini et al., 2023). Patients were supine on a stretcher with 0° of trunk flexion and knee fully extended, and each measure was repeated three times.

VI and rectus femoris (RF) thicknesses were measured bilaterally with the probe orientated longitudinally at 50% of the total distance from the superior patellar border towards the anterior superior iliac spine measured using metric plastic tape to represent the middle portion of the upper leg (Figure 1). Vastus medialis (VM) thickness was measured bilaterally from 4 cm medially and superior to the medial patella border (Figure 1). VL thickness was measured at 4 cm laterally from the VI and RF measure sites. Low-intense isometric contraction and palpation were performed before the acquisition to check VL location (Figure 1). The probe was perpendicular to the skin and aligned toward the muscle fibers. Special attention was given to obtaining parallel aponeurosis images between VI and RF and fiber differentiation between RF and VL (Figure 1). The Quadriceps portion thicknesses were vertically measured between muscles aponeurosis using the screen distance measure option of the ultrasound device (Philips Ultrasound, Inc., The Netherlands). To improve the reproducibility, we used standardized protocols for probe localization and orientation, such as those described previously, and also considered anatomical landmarks and probe localization in each patient. The Quadriceps thickness has been reported to have high reliability, ranging between 0.76 and 0.95 (Abiko et al., 2022).

**Figure 1.**
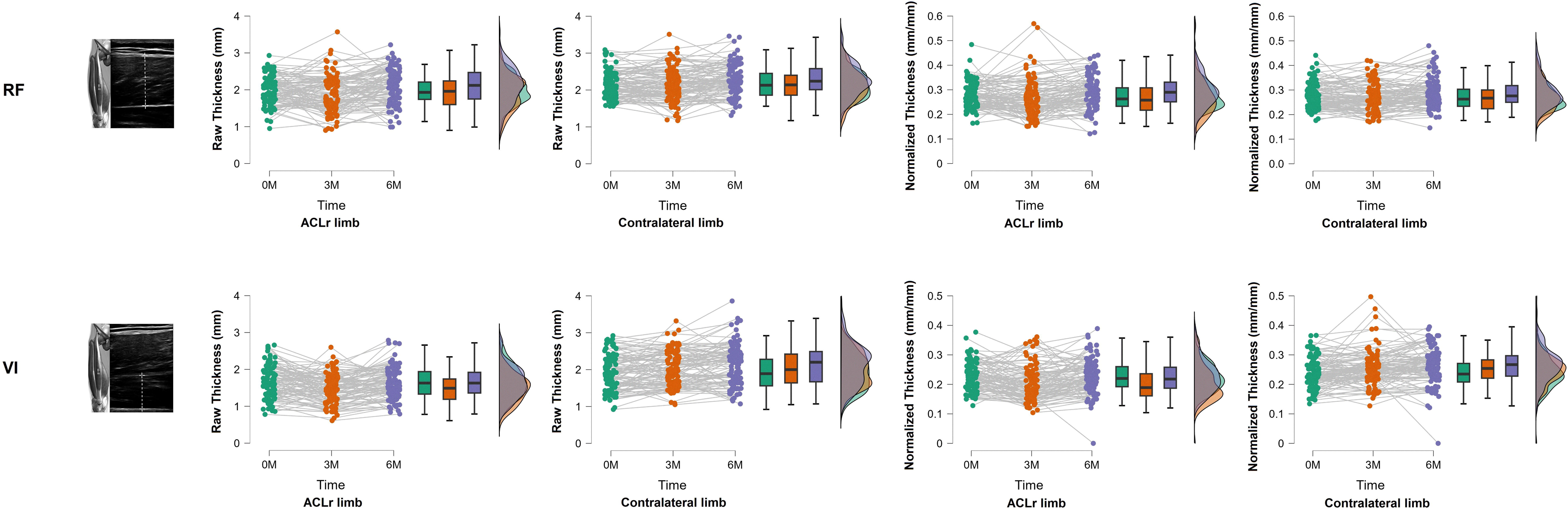
Time course thickness adaptation of quadriceps after an Anterior Cruciate Ligament reconstruction for Rectus Femoris (RF) and Vastus Intermedius (VI). The upper row of figures shows the ultrasonography probe location on RF and the raw and normalized outputs for the limb with Anterior Cruciate Ligament reconstruction (ACLr) and contralateral. The bottom row of the figures shows the ultrasonography probe location on VI, and the raw and normalized outputs for the limb with ACLr and contralateral.

**Figure 2.**
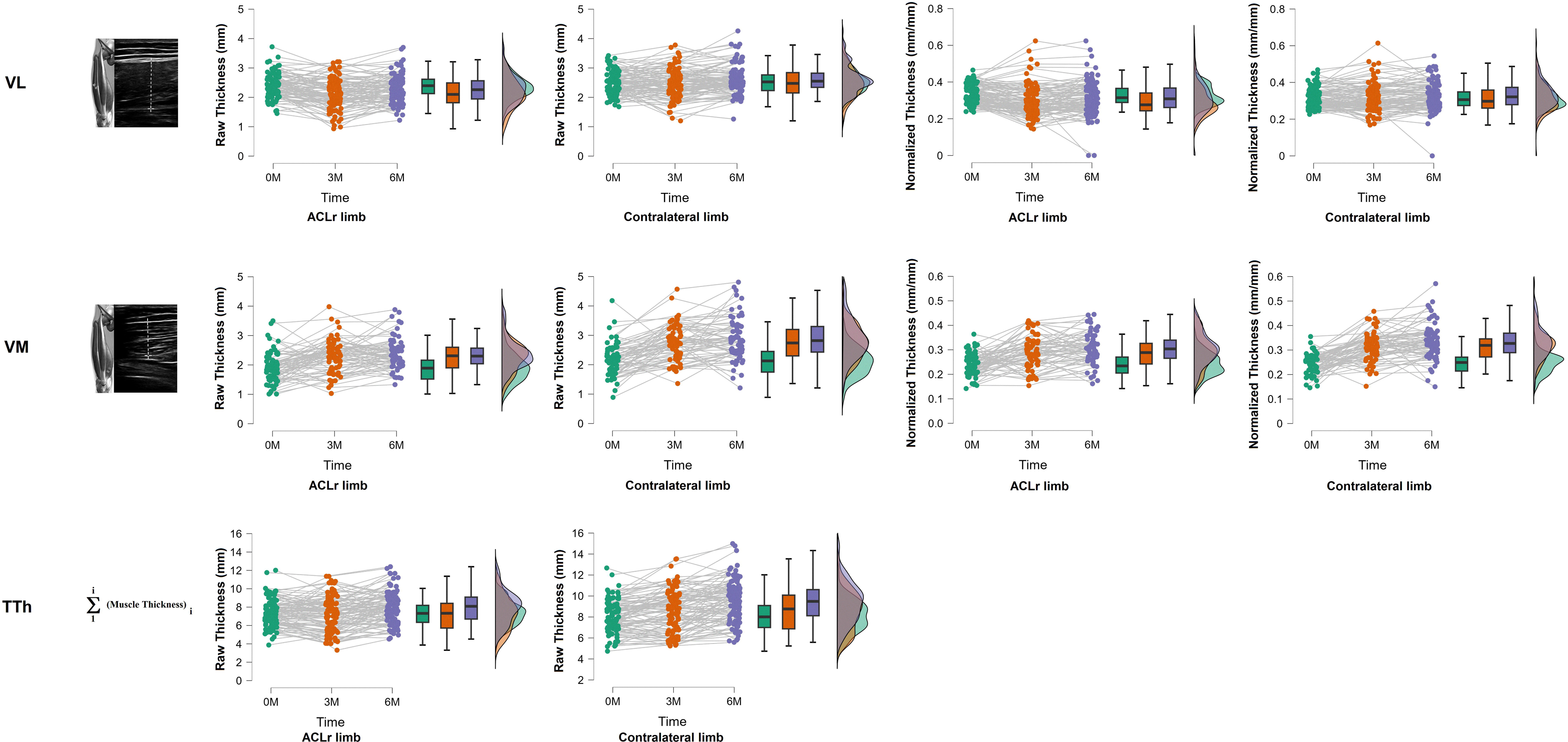
Time course thickness adaptation of quadriceps after an Anterior Cruciate Ligament reconstruction for Vastus Lateralis (VL), Vasutus Medialis (VM) and Total Thickness (TTh). The upper row of figures shows the ultrasonography probe location on VL and the raw and normalized outputs for the limb with Anterior Cruciate Ligament reconstruction (ACLr) and contralateral. The middle row of figures shows the ultrasonography probe location on VM, and the raw and normalized outputs for the limb with ACLr and contralateral. The bottom row of the figures shows the Total Thickness (TTh) estimation and the raw outputs for the limb with ACLr and contralateral.

Ultrasonographic variables were raw Quadriceps thickness measurements in mm units and normalized thickness measurements mm mm^-1^. Each portion of the Quadriceps was normalized to the total Quadriceps thickness estimated as the sum of VI, VL, VM, and RF thicknesses.

#### 2.5. Strength assessment

Isometric knee extension strength was evaluated. Strength was assessed through isometric tests to reduce patient risks in the first months of rehabilitation. Isometric Quadriceps strength was measured with the patients seated with the trunk in an upright posture and hip flexed at 90° and knee flexed at 90° in a custom chair instrumented with a S-Beam load cell (Rock Exotica LLC, USA, resolution ±2%, dynamic range ±1 kN to ±2 kN) attached at the ankle level (De la Fuente et al., 2023). The force signals were sampled at 500 Hz and measured in kgf through the Bluetooth Enforcer App. (Version 1.11488, Rock Exotica LLC, USA) using an iPad (Second generation, Apple Inc., USA). Patients were requested to complete two trials of maximal voluntary isometric Quadriceps contraction lasting 3 s each, with 1 min rest between trials, and the higher peak was considered for the statistical analyses. Patient was previously familiarized around 5 minuts regarding the test. The peak force was obtained from low-pass filtered signals with a Butterworth filter and a cutoff frequency of 50 Hz. The peak force value was converted from kgf to Nm units by multiplying by a constant of 9.81 m.s^-2^. Knee extension strength was normalized to the individual body mass.

#### 2.6. Complementary data collection

Thigh circumference was bilaterally measured using a metric plastic tape graded in millimeters at 5 and 20 cm above the patella’s middle portion and superior border (Shaw et al., 2004; Soderberg et al., 1996). Three measurements were made using the mean measure. After the surgery, the same physical therapist performed the measurements at 0M, 3M, and 6M.

#### 2.7. Data analysis

Quadriceps strength, normalized Quadriceps strength, thigh circumference at 5 cm and 20 cm, RF, VI, VL, VM and total thickness, normalized RF, VI, VL, and VM, are described considering the mean and standard deviation after confirming normality distribution (Kolmogorov-Smirnov test) and homoscedasticity (Levene’s test) principles. Two-way repeated measures ANOVA for factors time (0M, 3M, and 6M) and group (ACLr and CON group) with Bonferroni correction for multiple comparisons were performed to explore group differences and changes during the time course after surgery with time*group <0.05. Otherwise, a one-way repeated measure ANOVA with Bonferroni post hoc was used to verify time effects.

The Intraclass Correlation Coefficient (ICC) was obtained for *k* averaged measurements for a fixed evaluator as ICC_2,k_, where the random and systematic errors are considered in the ICC estimation (Weir, 2005). The mean squared of participants, error, and ICC_2,k_ calculation trials were obtained from a repeated measurement ANOVA design (Weir, 2005).

Finally, we performed a mediation analysis as part of structured equation models for each Quadriceps thickness, total thickness as a mediator, and Quadriceps strength as the output model. We considered the whole data as a time series for factors, mediators, and output. The model evaluates the indirect effect of thickness proportions involved in Quadriceps strength versus the direct effect of each muscle thickness without considering their effect on Quadriceps strength. All tests considered an α set at 5%, and all estimations were performed through the open-source software JASP 0.18.3 (University of Amsterdam, The Netherlands).

### 3. Results

Raw and normalized thicknesses results are sumarized in Table 1, Figure 1, and supplementary material.

The raw Quadriceps strength and Thigh circumferences is sumarized in Table 1 and Figure 3.

**Figure 3.**
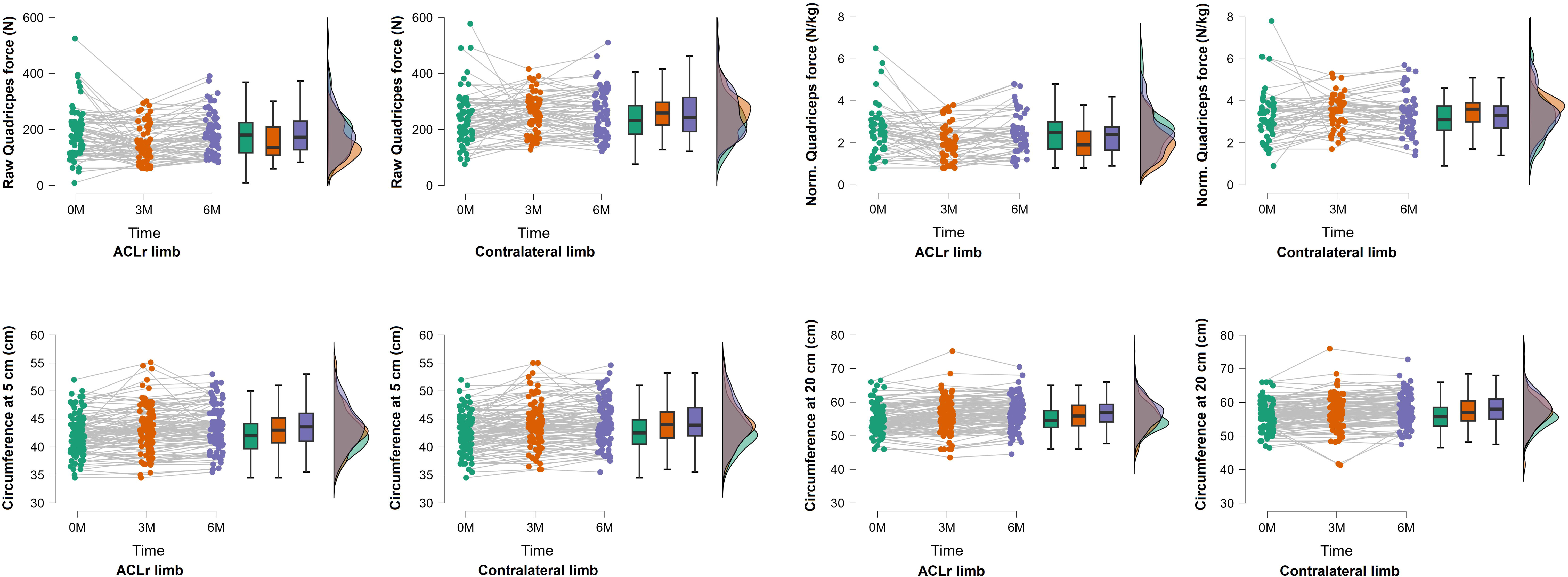
Quadriceps strength and knee circumference within 6 months after Anterior Cruciate Ligament reconstruction. The upper row of figures shows the raw and normalized quadriceps strength for the limb with Anterior Cruciate Ligament reconstruction (ACLr) and contralateral. The bottom row of the figures shows the raw and normalized circumferences for the limb with ACLr and contralateral.

Table 2 and Figure 4 summarize the mediation analysis of the normalized time course (time series) muscle thickness of ACLr during the six-month postoperative.

**Figure 4.**
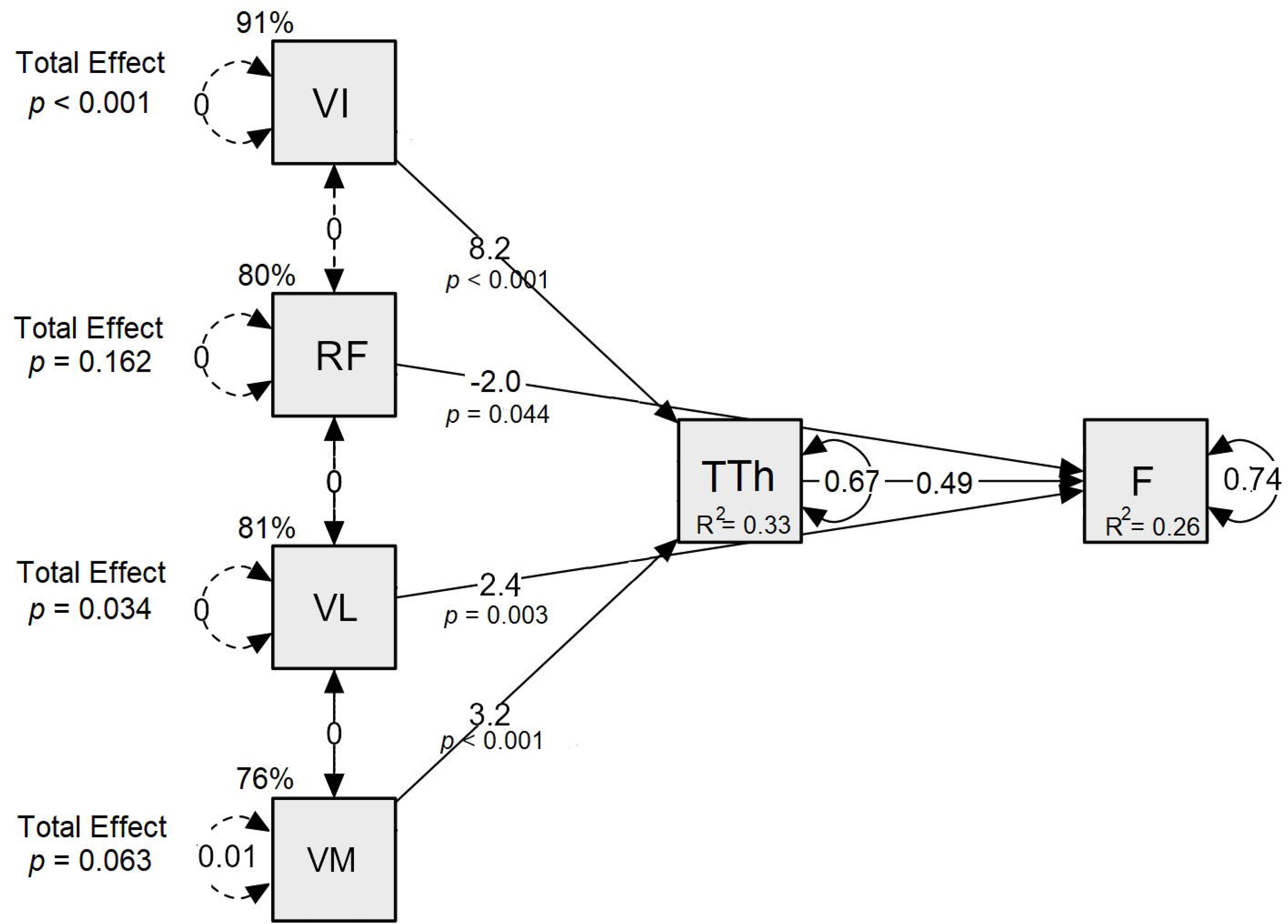
Mediation analysis on the time course of the quadriceps muscle strength (F). The vastus intermedius (VI) and medialis (VM) Thickness have an indirect effect on muscle Force (F) through the Total Thickness of the quadriceps (Th). The rectus femoris (RF) and vastus lateralis Thickness (VL) have a direct effect on muscle Force (F) following an Anterior Cruciate Ligament reconstruction within the first six months. Betas coefficients are described across the path, the variance of each latent factor as a double arrow, Covariance as straight vertical lines, the determinant coefficient as R-square, and the significant effect as a percentage.

### 4. Discussion

Our study provides insights into the thickness adaptation of Quadriceps muscle heads up to 6 months post-ACLr, demonstrating that VI and VL thickness adaptation determines the Quadriceps strength weakness following an ACLr. We found heterogenic changes across the four Quadriceps muscle heads. From 0 to 3 months, the decrease in VI and VL thicknesses determined the highest Quadriceps strength weakness. From 3 to 6 months, the increase in Quadriceps strength was determined by the increase in VI and VL thicknesses and total Quadriceps thickness. Up to 3 months, VM and RF showed a pattern of increasing thickness that aligns with changes in circumference measurements. However, six months after ACLr, Quadriceps strength, circumference at 5 cm, VI, VL, VM, and total thickness were not fully recovered compared to the CON limb. VI did not recover their Quadriceps proportion, and VI and VL did not recover to the pre-surgery conditions.

A novelty of our study was to identify a critical period within the first three months after ACLr when a peak of VI and VL atrophy was observed in the ACLr limb. The atrophy is likely to explain the ACLr Quadriceps strength deficits, which mediation analysis confirmed. Our findings agree with the smaller total Quadriceps, VI, and VL volumes and cross-sectional MRI measurement trends in the ACLr limb without relevant atrophy of other Quadriceps portions at 3.2 ± 2.0 months post-ACLr (Williams et al., 2005). In the first three months post-surgery, skeletal muscle repair and remodeling are suggested to be downregulated, causing apoptosis and muscle fiber atrophy (Parstorfer et al., 2021). As a result, there is a reduction in the sarcomere population, altering muscle mechanics contraction (Thomas et al., 2016). Furthermore, dysregulation of myokines [5] and muscle inhibition signaling may start as early as 72 hours post-surgery [7]. We argue that these physiological changes affect both VI and VL more than the other Quadriceps muscle heads, explaining its persistent impairments. The mediation analysis suggests that Quadriceps strength is mainly influenced by the thickness of VI, followed by VL and VM, while the RF thickness responds inversely (hypertrophy) over six-month post surgery.

The VI atrophy may precede the dysregulation of Quadriceps action (Kuenze et al., 2016) since VI thickness is considered a strong predictor of knee extensor strength (Ando et al., 2015; Kuenze et al., 2016) and the VI activation plays a stabilizing role in the knee (Alessandro et al., 2020; Mazzoli et al., 2018; Saito and Akima, 2015). The stabilizing function of the VI is observed within the first 50% of the stance phase in rat locomotion, showing different activation patterns compared to the other Quadriceps heads (Alessandro et al., 2020). In stroke survivors’, premature and prolonged VI activation is associated with a stiffer knee during gait (Mazzoli et al., 2018), smoother knee joint motion during hip flexion (Ando et al., 2015; Kuenze et al., 2016), and during Quadriceps contraction at >60% of the maximal voluntary isometric contraction VI shows an inverse activation pattern compared to the other Quadriceps portions (Akima and Saito, 2013). Therefore, the persistent atrophy pattern found is likely to predispose changes in the synergy of Quadriceps heads for effective knee extension force. This condition is not exclusively caused by VL dysfunction as previously was suggested for persistent Quadriceps weakness (Nuccio et al., 2021). In the same manner, it is expected knee stabilization impairements and induction of pathological stress over the patellar tendon by asymmetrical tractions. The reduced thickness of VI and VL appears to be compensated by the increased RF and VM capacity, as was evidenced by their increased thickness throughout the ACLr time course in our study.

The time course for VM and RF showed a progressive thickness increase in the ACLr and CON limb after six months post-surgery. These patterns were only in coherence with the patterns showed by the circumferences at 5 and 20 cm over the knee. The progressive thickness increase of VM and RF suggests a hypertrophy adaptation, in contrast to the observed in the VI and VL. Unfortunately, the hypertrophy of VM and hypotrophy of VL may lead to disbalances in the mediolateral knee force, which agrees with patients who develop early patellofemoral pain after ACLr (Ong et al., 2023). Our findings for VM and RF also suggest that the synergy actions between Quadriceps portions can change after six months post-surgery.

Although ACLr studies frequently focus on VM or VL due to patellofemoral pain (Ong et al., 2023) or muscle inhibition (Nuccio et al., 2021), early VI atrophy has not been considered during ACLr rehabilitation within the first three months post surgery. Therefore, a more comprehensive understanding of vastii atrophy and its therapeutics, such as early rehabilitation, early physiological stimulus (De la Fuente et al., 2024), early feedback methods (De la Fuente et al., 2020), or muscle strengthening aiming for each Quadriceps portion (Narici et al., 1996) could lead to better clinical outcomes as observed in other pathologies (C. De la Fuente et al., 2016; De la Fuente et al., 2024, 2020, 2018; C. I. De la Fuente et al., 2016). Also, the observed deficit in the total Quadriceps thickness compared to the CON limb establishes a risk factor for lower limb injury (Koźlenia et al., 2022) and agrees with morphological impairments as the primary source of strength deficits following ACLr (Birchmeier et al., 2020; Krishnan and Williams, 2011; Norte et al., 2018; Thomas et al., 2016; Tim-Yun Ong et al., 2022). In addition, we call attention to the low sensitivity of thigh circumference measurement to known specific Quadriceps portion thickness changes following an ACLr. Finally, assessing the time course of muscle thickness until six months is not trivial because it is a standard period to begin with sports movements (Williams et al., 2005) and would be a period when Quadriceps activation failure can be more evident (De la Fuente et al., 2022b).

The main limitations of our study include the different training levels of the participants before the surgery (no athletes were included), daily routine during the rehabilitation (because of the inherent differences in life routines between participants), the type of graft (not all patients required the same technique), sex (inherent to physiological sex differences), undesired dropout of patients (a personal decision of patients or incapacity to assist by external factors which were not considered for repeated measurement methods) that would have added more data variability, and the low ICC values for VM measurements. Therefore, readers need to interpret the study considering these limitations.

### 5. Conclusions

Quadriceps thickness is not recovered up to six months following an ACLr surgery. Until the third month post-surgery, VI and VL are highly atrophied, compromising the Quadriceps strength and establishing the most critical rehabilitation stage. VI does not recover its Quadriceps proportion, and VI and VL do not recover pre-surgery conditions, which are the primary determinants of Quadriceps strength changes following ACLr. Finally, VI thickness may be a used as a biomarker predicting Quadriceps weakness following ACLr.

## Data Availability

All data produced in the present study are available upon reasonable request to the authors

## Acknowledgment

1. C. De la Fuente and A. de Andrade acknowledge Departamento de Esportes, Escola de Educaçao Física, Fisioterapía e Terapía Ocupacional, EEFFTO-UFMG, Universidade Federal do Minas Gerais, Belo Horizonte, MG, Brazil where C. De la Fuente is doing his voluntary (non-funded) postdoctorate in functional and time-series statistics. The authors thank C. Fleckenstein and M. Soldan for the data collection. Authors C. De la Fuente, R. Yañez, R., and Silvestre thank *Sociedad Chile de Traumatologia y Ortopedia* for the *Concurso de Ligamento Cruzado Anterior 2024* funding No. 059. E. Martinez-Valdes is supported by an early career research fellowship grant from Orthopaedic Research UK (ORUK ref: 574).

## References

Abiko, T., Ohmae, K., Murata, S., Shiraiwa, K., Horie, J., 2022. Reliability of muscle thickness and echo intensity measurements of the quadriceps: A novice examiner. J Bodyw Mov Ther 31, 164–168. 10.1016/j.jbmt.2022.03.004

Agarwal, S., Jain, H., Raichandani, K., Godara, J., Choudhary, S., 2021. Early Quadriceps Wasting after Anterior Cruciate Ligament Reconstruction in Young Adults: A Prospective Study. JCDR. 10.7860/JCDR/2021/47254.14769

Akima, H., Saito, A., 2013. Inverse activation between the deeper vastus intermedius and superficial muscles in the quadriceps during dynamic knee extensions. Muscle Nerve 47, 682–690. 10.1002/mus.23647

Alessandro, C., Barroso, F.O., Prashara, A., Tentler, D.P., Yeh, H.-Y., Tresch, M.C., 2020. Coordination amongst quadriceps muscles suggests neural regulation of internal joint stresses, not simplification of task performance. Proc Natl Acad Sci U S A 117, 8135–8142. 10.1073/pnas.1916578117

Ando, R., Saito, A., Umemura, Y., Akima, H., 2015. Local architecture of the vastus intermedius is a better predictor of knee extension force than that of the other quadriceps femoris muscle heads. Clin Physiol Funct Imaging 35, 376–382. 10.1111/cpf.12173

Birchmeier, T., Lisee, C., Kane, K., Brazier, B., Triplett, A., Kuenze, C., 2020. Quadriceps Muscle Size Following ACL Injury and Reconstruction: A Systematic Review. J Orthop Res 38, 598–608. 10.1002/jor.24489

Boccia, G., Martinez-Valdes, E., Negro, F., Rainoldi, A., Falla, D., 2019. Motor unit discharge rate and the estimated synaptic input to the vasti muscles is higher in open compared with closed kinetic chain exercise. J Appl Physiol (1985) 127, 950–958. 10.1152/japplphysiol.00310.2019

Bouillard, K., Jubeau, M., Nordez, A., Hug, F., 2014. Effect of vastus lateralis fatigue on load sharing between quadriceps femoris muscles during isometric knee extensions. J Neurophysiol 111, 768–776. 10.1152/jn.00595.2013

Caracciolo, G., Yáñez, R., Silvestre, R., De la Fuente, C., Zamorano, H., Ossio, A., Strömbäck, L., Abusleme, S., P. Carpes, F., 2021. Intraoperative pivot-shift accelerometry combined with anesthesia improves the measure of rotatory knee instability in anterior cruciate ligament injury. J Exp Orthop 8, 80. 10.1186/s40634-021-00396-1

Cobian, D.G., Knurr, K.A., Joachim, M.R., Bednarek, A.L., Broderick, A.M., Heiderscheit, B.C., 2024. Does It Matter? Isometric or Isokinetic Assessment of Quadriceps Strength Symmetry 9 Months After ACLR in Collegiate Athletes. Sports Health 19417381241247819. 10.1177/19417381241247819

Criss, C.R., Lepley, A.S., Onate, J.A., Clark, B.C., Simon, J.E., France, C.R., Grooms, D.R., 2023. Brain activity associated with quadriceps strength deficits after anterior cruciate ligament reconstruction. Sci Rep 13, 8043. 10.1038/s41598-023-34260-2

Czaplicki, A., Jarocka, M., Walawski, J., 2015. Isokinetic Identification of Knee Joint Torques before and after Anterior Cruciate Ligament Reconstruction. PLoS One 10, e0144283. 10.1371/journal.pone.0144283

De la Fuente, C., Cruz-Montencinos, C., la Fuente, C.D., y Lillo, R.P., Chamorro, C., Henriquez, H., 2018. Early Short-Term Recovery of Single-Leg Heel Rise and ATRS After Achilles Tenorrhaphy: Cluster Analysis [WWW Document]. Asian Journal of Sports Medicine. 10.5812/asjsm.67661

De la Fuente, C., Neira, A., Torres, G., Silvestre, R., Roby, M., Yañez, R., Herrera, S., Martabit, V., McKay, I., Carpes, F.P., 2022a. Effects of Elbow Crutch Locomotion on Gluteus Medius Activation During Stair Ascending. Front Bioeng Biotechnol 10, 890004. 10.3389/fbioe.2022.890004

De la Fuente, C., Peña y Lillo, R., Carreño, G., Marambio, H., 2016. Prospective randomized clinical trial of aggressive rehabilitation after acute Achilles tendon ruptures repaired with Dresden technique. Foot (Edinb) 26, 15–22. 10.1016/j.foot.2015.10.003

De la Fuente, C., Silvestre, R., Baechler, P., Gemigniani, A., Grunewaldt, K., Vassiliu, M., Wodehouse, V., Delgado, M., Carpes, F.P., 2020. Intrasession Real-time Ultrasonography Feedback Improves the Quality of Transverse Abdominis Contraction. J Manipulative Physiol Ther. 10.1016/j.jmpt.2019.10.017

De la Fuente, C., Silvestre, R., Botello, J., Neira, A., Soldan, M., Carpes, F.P., 2024. Unique case study: Impact of single-session neuromuscular biofeedback on motor unit properties following 12 days of Achilles tendon surgical repair. Physiol Rep 12, e15868. 10.14814/phy2.15868

De la Fuente, C., Silvestre, R., Yañez, R., Roby, M., Soldán, M., Ferrada, W., Carpes, F.P., 2023. Preseason multiple biomechanics testing and dimension reduction for injury risk surveillance in elite female soccer athletes: short-communication. Sci Med Footb 7, 183–188. 10.1080/24733938.2022.2075558

De la Fuente, C., Stoelben, K.J.V., Silvestre, R., Yañez, R., Cheyre, J., Guadagnin, E.C., Carpes, F.P., 2022b. Steadiness training improves the quadriceps strength and self-reported outcomes in persistent quadriceps weakness following nine months of anterior cruciate ligament reconstruction and failed conventional physiotherapy. Clin Biomech (Bristol, Avon) 92, 105585. 10.1016/j.clinbiomech.2022.105585

De la Fuente, C.I., Lillo, R.P.Y., Ramirez-Campillo, R., Ortega-Auriol, P., Delgado, M., Alvarez-Ruf, J., Carreño, G., 2016. Medial Gastrocnemius Myotendinous Junction Displacement and Plantar-Flexion Strength in Patients Treated With Immediate Rehabilitation After Achilles Tendon Repair. J Athl Train 51, 1013–1021. 10.4085/1062-6050-51.12.23

Fukunaga, T., Johnson, C.D., Nicholas, S.J., McHugh, M.P., 2019. Muscle hypotrophy, not inhibition, is responsible for quadriceps weakness during rehabilitation after anterior cruciate ligament reconstruction. Knee Surg Sports Traumatol Arthrosc 27, 573–579. 10.1007/s00167-018-5166-1

Grapar Žargi, T., Drobnič, M., Vauhnik, R., Koder, J., Kacin, A., 2017. Factors predicting quadriceps femoris muscle atrophy during the first 12weeks following anterior cruciate ligament reconstruction. Knee 24, 319–328. 10.1016/j.knee.2016.11.003

Harput, G., Demirci, S., Soylu, A.R., Bayrakci Tunay, V., 2022. Association between quadriceps muscle thickness and knee function in anterior cruciate ligament reconstructed athletes: a cross-sectional study. Physiother Theory Pract 1–9. 10.1080/09593985.2022.2068096

Hennebry, A., Oldham, J., Shavlakadze, T., Grounds, M.D., Sheard, P., Fiorotto, M.L., Falconer, S., Smith, H.K., Berry, C., Jeanplong, F., Bracegirdle, J., Matthews, K., Nicholas, G., Senna-Salerno, M., Watson, T., McMahon, C.D., 2017. IGF1 stimulates greater muscle hypertrophy in the absence of myostatin in male mice. J Endocrinol 234, 187–200. 10.1530/JOE-17-0032

Irrgang, J.J., Anderson, A.F., Boland, A.L., Harner, C.D., Kurosaka, M., Neyret, P., Richmond, J.C., Shelborne, K.D., 2001. Development and validation of the international knee documentation committee subjective knee form. Am J Sports Med 29, 600–613. 10.1177/03635465010290051301

Koźlenia, D., Struzik, A., Domaradzki, J., 2022. Force, Power, and Morphology Asymmetries as Injury Risk Factors in Physically Active Men and Women. Symmetry 14, 787. 10.3390/sym14040787

Krishnan, C., Williams, G.N., 2011. Factors explaining chronic knee extensor strength deficits after ACL reconstruction. J Orthop Res 29, 633–640. 10.1002/jor.21316

Kuenze, C.M., Blemker, S.S., Hart, J.M., 2016. Quadriceps function relates to muscle size following ACL reconstruction. J Orthop Res 34, 1656–1662. 10.1002/jor.23166

Kuenze, C.M., Hertel, J., Weltman, A., Diduch, D., Saliba, S.A., Hart, J.M., 2015. Persistent Neuromuscular and Corticomotor Quadriceps Asymmetry After Anterior Cruciate Ligament Reconstruction. J Athl Train 50, 303–312. 10.4085/1062-6050-49.5.06

Lanferdini, F.J., Diefenthaeler, F., Ávila, A.G., Moro, A.R.P., van der Zwaard, S., Vaz, M.A., 2023. Quadriceps Muscle Morphology Is an Important Determinant of Maximal Isometric and Crank Torques of Cyclists. Sports (Basel) 11, 22. 10.3390/sports11020022

Lee, J.-H., Cheon, S., Jun, H.-P., Huang, Y.-L., Chang, E., 2020. Bilateral Comparisons of Quadriceps Thickness after Anterior Cruciate Ligament Reconstruction. Medicina (Kaunas) 56, 335. 10.3390/medicina56070335

Mazzoli, D., Giannotti, E., Manca, M., Longhi, M., Prati, P., Cosma, M., Ferraresi, G., Morelli, M., Zerbinati, P., Masiero, S., Merlo, A., 2018. Electromyographic activity of the vastus intermedius muscle in patients with stiff-knee gait after stroke. A retrospective observational study. Gait Posture 60, 273–278. 10.1016/j.gaitpost.2017.07.002

Narici, M.V., Hoppeler, H., Kayser, B., Landoni, L., Claassen, H., Gavardi, C., Conti, M., Cerretelli, P., 1996. Human quadriceps cross-sectional area, torque and neural activation during 6 months strength training. Acta Physiol Scand 157, 175–186. 10.1046/j.1365-201X.1996.483230000.x

Norte, G.E., Knaus, K.R., Kuenze, C., Handsfield, G.G., Meyer, C.H., Blemker, S.S., Hart, J.M., 2018. MRI-Based Assessment of Lower-Extremity Muscle Volumes in Patients Before and After ACL Reconstruction. J Sport Rehabil 27, 201–212. 10.1123/jsr.2016-0141

Nuccio, S., Del Vecchio, A., Casolo, A., Labanca, L., Rocchi, J.E., Felici, F., Macaluso, A., Mariani, P.P., Falla, D., Farina, D., Sbriccoli, P., 2021. Deficit in knee extension strength following anterior cruciate ligament reconstruction is explained by a reduced neural drive to the vasti muscles. J Physiol 599, 5103–5120. 10.1113/JP282014

Ong, M.T., Chi-wai Man, G., He, X., Yu, M., Lau, L.C., Qiu, J., Wang, Q., Ho-pak Liu, J., Chi-yin Choi, B., Ng, J.P., Shu-hang Yung, P., 2023. Assessments of early patellofemoral joint osteoarthritis features after anterior cruciate ligament reconstruction: a cross-sectional study. BMC Musculoskeletal Disorders 24, 510. 10.1186/s12891-023-06639-9

Parstorfer, M., Profit, F., Weiberg, N., Wehrstein, M., Barié, A., Friedmann-Bette, B., 2021. Increased satellite cell apoptosis in vastus lateralis muscle after anterior cruciate ligament reconstruction. J Rehabil Med 53, jrm00153. 10.2340/16501977-2794

Pietrosimone, B., Lepley, A.S., Kuenze, C., Harkey, M.S., Hart, J.M., Blackburn, J.T., Norte, G., 2022. Arthrogenic Muscle Inhibition Following Anterior Cruciate Ligament Injury. J Sport Rehabil 31, 694–706. 10.1123/jsr.2021-0128

Pottkotter, K.A., Di Stasi, S.L., Schmitt, L.C., Magnussen, R.A., Paterno, M.V., Flanigan, D.C., Kaeding, C.C., Hewett, T.E., 2020. Timeline of gains in quadriceps strength symmetry and patient-reported function early after ACL reconstruction. Int J Sports Phys Ther 15, 995– 1005. 10.26603/ijspt20200995

Saito, A., Akima, H., 2015. Neuromuscular Activation of the Vastus Intermedius Muscle during Isometric Hip Flexion. PLoS One 10, e0141146. 10.1371/journal.pone.0141146

San Martín-Mohr, C., Cristi-Sánchez, I., Pincheira, P.A., Reyes, A., Berral, F.J., Oyarzo, C., 2018. Knee sensorimotor control following anterior cruciate ligament reconstruction: A comparison between reconstruction techniques. PLoS One 13, e0205658. 10.1371/journal.pone.0205658

Shaw, T., Chipchase, L.S., Williams, M.T., 2004. A users guide to outcome measurement following ACL reconstruction. Physical Therapy in Sport 5, 57–67. 10.1016/j.ptsp.2003.11.007

Soderberg, G.L., Ballantyne, B.T., Kestel, L.L., 1996. Reliability of lower extremity girth measurements after anterior cruciate ligament reconstruction. Physiother Res Int 1, 7–16. 10.1002/pri.43

Sonnery-Cottet, B., Saithna, A., Quelard, B., Daggett, M., Borade, A., Ouanezar, H., Thaunat, M., Blakeney, W.G., 2019. Arthrogenic muscle inhibition after ACL reconstruction: a scoping review of the efficacy of interventions. Br J Sports Med 53, 289–298. 10.1136/bjsports-2017-098401

Thomas, A.C., Wojtys, E.M., Brandon, C., Palmieri-Smith, R.M., 2016. Muscle atrophy contributes to quadriceps weakness after anterior cruciate ligament reconstruction. J Sci Med Sport 19, 7–11. 10.1016/j.jsams.2014.12.009

Tim-Yun Ong, M., Fu, S.-C., Mok, S.-W., Franco-Obregón, A., Lok-Sze Yam, S., Shu-Hang Yung, P., 2022. Persistent quadriceps muscle atrophy after anterior cruciate ligament reconstruction is associated with alterations in exercise-induced myokine production. Asia Pac J Sports Med Arthrosc Rehabil Technol 29, 35–42. 10.1016/j.asmart.2022.05.001

Weir, J.P., 2005. Quantifying test-retest reliability using the intraclass correlation coefficient and the SEM. J Strength Cond Res 19, 231–240. 10.1519/15184.1

Williams, G.N., Buchanan, T.S., Barrance, P.J., Axe, M.J., Snyder-Mackler, L., 2005. Quadriceps weakness, atrophy, and activation failure in predicted noncopers after anterior cruciate ligament injury. Am J Sports Med 33, 402–407. 10.1177/0363546504268042

Yañez-Diaz, R., Roby, M., Silvestre, R., Zamorano, H., Vergara, F., Sandoval, C., Neira, A., Yañez-Rojo, C., De la Fuente, C., 2023. Multiclass Support Vector Machine improves the Pivot-shift grading from Gerdy’s acceleration resultant prior to the acute Anterior Cruciate Ligament surgery. Injury S0020-1383(23)00271–1. 10.1016/j.injury.2023.03.020

Yang, J.-H., Eun, S.-P., Park, D.-H., Kwak, H.-B., Chang, E., 2019. The Effects of Anterior Cruciate Ligament Reconstruction on Individual Quadriceps Muscle Thickness and Circulating Biomarkers. Int J Environ Res Public Health 16, 4895. 10.3390/ijerph16244895

