## Supplementary material for "Time course of Quadriceps thickness changes over six months post-anterior cruciate ligament reconstruction: unveiling critical impairments in vastii muscles": Table 1

**Table 1.** Time course outcomes of the study.

| Measurements | ACLr limb | | | | Non-injured limb | | | | 2-way RM ANOVA | 1-way RM ANOVA |
|---|---|---|---|---|---|---|---|---|---|---|
| | Pre-surgery | Month 3 | Month 6 | $ICC_{2,k}$ | Pre-surgery | Month 3 | Month 6 | $ICC_{2,k}$ | Time*group, $\eta_p^2$ | Time effect, $\eta_p^2$ |
| ***Circumference:*** | | | | | | | | | | |
| Circumference at 5cm (cm) | 42.3 ± 3.5[ʁ,ǰ] | 43.2 ± 4.1[ʁ,Λ] | 43.7 ± 3.8[ǰ,и] | 0.92[þ] | 42.5 ± 3.4[ʁ,ǰ] | 44.1 ± 4.0[ʁ,Λ] | 44.4 ± 3.9[ǰ,и] | 0.86[ϱ] | *p* = 0.010, 0.008 | |
| Circumference at 20 cm (cm) | 54.8 ± 4.2[ʒ,ʃ] | 55.1 ± 4.1[ʒ,Ч] | 56.8 ± 4.5[ʃ,Ч] | 0.92[þ] | 56.0 ± 4.0[ʁ,ʃ] | 57.2 ± 5.0[ʁЧ] | 58.0 ± 4.5[,ʃ,Ч] | 0.90[þ] | *p* = 0.058, 0.003 | $p_{ACLr}$ < 0.001, 0.196<br>$p_{CON}$ < 0.001, 0.257 |
| ***Raw Thickness:*** | | | | | | | | | | |
| RF Thickness (cm) | 1.98 ± 0.38 | 1.93 ± 0.49[б] | 2.11 ± 0.48[б] | 0.69[Ш] | 2.16 ± 0.37 | 2.14 ± 0.45[б] | 2.28 ± 0.43[б] | 0.74[Ш] | *p* = 0.564, < 0.001 | $p_{ACLr}$ < 0.001, 0.074 |
| VI Thickness (cm) | 1.66 ± 0.43[ʁ,ɐ] | 1.48 ± 0.40[ʁ,б,Λ] | 1.64 ± 0.43[б,Ɵ] | 0.68[Ш] | 1.92 ± 0.48[ʃ,ɐ] | 2.03 ± 0.48[Λ] | 2.13 ± 0.55[ʃ,Ɵ] | 0.80[ϱ] | *p* < 0.001, 0.028 | $p_{CON}$ < 0.001, 0.064 |
| VL Thickness (cm) | 2.39 ± 0.40[ʁ, ȣ] | 2.12 ± 0.51[ʁ,Ч, Λ] | 2.29 ± 0.48[Ч,Ɵ] | 0.70[Ш] | 2.51 ± 0.38[ȣ] | 2.47 ± 0.52[Ч, Λ] | 2.60 ± 0.51[Ч,Ɵ] | 0.80[ϱ] | *p* < 0.001, 0.025 | |
| VM Thickness (cm) | 1.91 ± 0.52[ʁ,ʃ,ɐ] | 2.29 ± 0.56[ʁ,Λ] | 2.37 ± 0.53[ʃ,Ɵ] | 0.53[Ш] | 2.13 ± 0.55[ʁ,ʃ,ɐ] | 2.75 ± 0.61[ʁ,Λ] | 2.89 ± 0.72[ʃ,Ɵ] | 0.48[ʅ] | *p* < 0.001, 0.015 | |
| Total thickness (cm) | 7.38 ± 1.49[ʃ,ɐ] | 7.21 ± 1.88[б,Λ] | 8.02 ± 1.76[ʃ,б,Ɵ] | 0.74[Ш] | 8.06 ± 1.55[ʒ,ʃ,ɐ] | 8.61 ± 1.97[ʒ,б,Λ] | 9.45 ± 1.98[ʃ,б,Ɵ] | 0.75[Ш] | *p* < 0.001, 0.020 | |
| ***Normalized Thickness:*** | | | | | | | | | | |
| RF Thickness (%) | 27.3± 5.3 | 26.8± 7.7[Ч] | 29.1± 6.5[Ч] | 0.75[ϱ] | 27.3± 5.1 | 27.1± 5.8[Ч] | 28.9± 6.2[Ч] | 0.83[ϱ] | *p* = 0.677, < 0.001 | $p_{ACLr}$ < 0.001, 0.074 |
| VI Thickness (%) | 22.6± 4.8[ʒ] | 20.4± 6.0[ʒ,Ч] | 22.4± 5.9[Ч] | 0.63[Ш] | 24.0± 5.1[ʂ] | 25.5± 6.0 | 26.4± 6.2[ʂ] | 0.68[Ш] | *p* < 0.001, 0.030 | $p_{CON}$ = 0.007, 0.048 |
| VL Thickness (%) | 32.9± 5.3[ʁ] | 29.5± 8.6[ʁ,ϖ] | 31.4± 9.4 | 0.74[Ш] | 31.7± 5.4 | 31.5± 7.8[ϖ] | 32.8± 7.6 | 0.81[ϱ] | *p* < 0.001, 0.023 | |
| VM Thickness (%) | 24.0± 4.6[ʁ,ʃ] | 29.0± 6.7[ʁ] | 30.3± 6.4[ʃ] | 0.28[ʅ] | 24.4± 4.6[ʁ,ʃ] | 31.4± 6.0[ʁ] | 33.3± 7.5[ʃ] | 0.11[ʅ] | *p* = 0.010, 0.009 | |
| ***Quadriceps strength:*** | | | | | | | | | *p* < 0.001, 0.027 | |
| Raw Quad. strength (N) | 188.1± 88.5[ʒ, ɐ] | 155.7± 64.5[ʒ,,Ч,, Λ] | 186.1± 73.1[Ч, Ɵ] | 0.74[Ш] | 238.4± 92.9[ɐ] | 254.9± 68.3[Λ] | 254.6± 81.5[Ɵ] | 0.77[ϱ] | *p* < 0.001, 0.033 | |
| Norm. Quad. strength (N $kg^{-1}$) | 2.59± 1.28[ʒ,ɐ] | 2.05± 0.85[ʒ, Λ] | 2.45±1.01[Ɵ] | 0.72[Ш] | 3.27± 1.27[ɐ] | 3.48± 0.81[Λ] | 3.37± 1.01[Ɵ] | 0.73[Ш] | | |

*RF = Rectus femoris; VI = Vastus intermedius; VL = Vastus lateralis; VM = Vastus medialis; Quad. = quadriceps; RM = Repeated measurements; ACLr = Anterior Cruciate Ligament reconstruction; ICC = Intraclass Coefficient Correlation; 2,k = Reliability calculated by taking an average of the k measurements considering random and systematic errors* [34].

[ȣ] *= statistical significance (p < 0.05) at 0M between limbs.*
[ɐ] *= statistical significance (p < 0.001) at 0M between limbs.*
[ϖ] *= statistical significance (p < 0.05) at 3M between limbs.*
[Λ] *= statistical significance (p < 0.001) at 3M between limbs.*
[и] *= statistical significance (p < 0.05) at 6M between limbs.*
[Ɵ] *= statistical significance (p < 0.001) at 6M between limbs.*
[ʒ] *= statistical significance (p < 0.05) between 0M and 3M within the limb.*
[ʁ] *= statistical significance (p < 0.001) between 0M and 3M within the limb.*
[ǰ] *= statistical significance (p < 0.05) between 0M and 6M within the limb.*
[ʃ] *= statistical significance (p < 0.001) between 0M and 6M within the limb.*
[Ч] *= statistical significance (p < 0.05) between 3M and 6M within the limb.*
[б] *= statistical significance (p < 0.001) between 3M and 6M within the limb.*
[ʅ] *= ICC ≥ 0.5,* [Ш]*= moderate ICC (0.50 to 0.75],* [ϱ] *= good ICC (0.75 to 0.90], and* [þ] *= perfect ICC ≥ 0.90.*
