## Supplementary material for "Time course of Quadriceps thickness changes over six months post-anterior cruciate ligament reconstruction: unveiling critical impairments in vastii muscles": Table 2

**Table 2.** Mediation analysis of the normalized time course muscle Thickness of ACLr.

| Effects: | Path | | | | β | SE | z-value | *p-value* |
|---|---|---|---|---|---|---|---|---|
| ***Direct effects:*** | | | | | | | | |
| Vastus Intermedius Thickness | → | ACLr Force | | | 0.396 | 0.973 | 0.407 | 0.684 |
| Rectus Femoris Thickness | → | ACLr Force | | | -1.998 | 0.994 | -2.010 | 0.044 |
| Vastus Lateralis Thickness | → | ACLr Force | | | 2.363 | 0.783 | 3.017 | 0.003 |
| Vastus Medialis Thickness | → | ACLr Force | | | -0.499 | 0.535 | -0.934 | 0.350 |
| ***Indirect effects:*** | | | | | | | | |
| Vastus Intermedius Thickness | → | Total Thickness | → | ACLr Force | 4.007 | 0.561 | 7.137 | <0.001 |
| Rectus Femoris Thickness | → | Total Thickness | → | ACLr Force | 0.487 | 0.430 | 1.133 | 0.257 |
| Vastus Lateralis Thickness | → | Total Thickness | → | ACLr Force | -0.550 | 0.349 | -1.575 | 0.115 |
| Vastus Medialis Thickness | → | Total Thickness | → | ACLr Force | 1.541 | 0.274 | 5.620 | <0.001 |
| ***Total effects:*** | | | | | | | | |
| Vastus Intermedius Thickness | → | ACLr Force | | | 4.403 | 0.972 | 4.532 | <0.001 |
| Rectus Femoris Thickness | → | ACLr Force | | | -1.511 | 1.080 | -1.399 | 0.162 |
| Vastus Lateralis Thickness | → | ACLr Force | | | 1.813 | 0.854 | 2.124 | 0.034 |
| Vastus Medialis Thickness | → | ACLr Force | | | 1.041 | 0.560 | 1.861 | 0.063 |

*SE = standard error.*
*ACLr = Anterior Cruciate Ligament reconstruction*
